# Integrative Modeling of SARS-CoV-2 Infection Dynamics to Inform COVID-19 Vaccination Strategies

**DOI:** 10.1101/2025.08.19.25333964

**Authors:** Rachel Waema, Charles Mwangi Kaumbutha, Titus Okello Orwa

## Abstract

Non-pharmaceutical interventions were very instrumental in the early phases of COVID-19 pandemic. Fortunately, the urgency to control the crisis prompted an accelerated vaccine development process, saving millions of lives globally. Despite these measures, cases of COVID-19 and other variants SARS-CoV-2 are still being reported in different countries and regions. We present a within-host mathematical model of SARS-CoV-2 infection that incorporates target cell dynamics, innate and adaptive immune responses, and vaccine interventions. Analytical results identify the basic reproduction number, *R*_0_, as the threshold for infection persistence. Simulations show that immune responses, both lytic and non-lytic, are critical in controlling viral replication, with vaccine-induced immunity further reducing viral load and protecting epithelial cells. Immune-boosting strategies and monoclonal antibody therapies targeting intracellular replication outperform entry-blocking interventions alone, while combination approaches yield the greatest reduction in peak viral load and fastest clearance. Timing is crucial: early vaccination or treatment maximizes benefits, whereas delays allow higher viral titers to persist. These results underscore the importance of early, multi-mechanism interventions, particularly for vulnerable populations such as the elderly and immunocompromised. The model offers a framework for evaluating treatment strategies and can be extended to incorporate pharmacokinetics/pharmacodynamics or patient-specific calibration for improved predictive accuracy.

## 1 Introduction

COVID-19 (Coronavirus disease 2019) is an infectious disease caused by the SARS-CoV-2 virus [11]. Most infections from coronaviruses are mild, however, previous SARS CoV and MERS-CoV epidemics generated more than 10000 cases collectively in the past two decades with MERS-CoV registering higher mortality rates of 37% against 10% for SARS CoV [13, 18]. COVID-19, which emerged from Wuhan, China in December 2019 [23] has caused over 760 million cases and 6.9 million deaths worldwide [40].

SARS CoV 2 belongs to a group of enveloped Corona viruses with a positive-sense, single stranded RNA and viral particles. The genomic sequence of SARS CoV-2 shares 80% similarity with SARS CoV and 50% with MERS-CoV [11]. The SARS-CoV-2 genome comprises of a capped and polyadenylated RNA. The single stranded RNA contains one large 5 prime open reading frame (ORF) and smaller ORFs. Proteolytic activity on the large ORF by viral proteases produce the non-structural proteins (NSP1-NSP16). The smaller ORFs code for structural proteins: spike (S), envelope (E), membrane (M) and nucleocapsid (N) and other polypeptides. Collectively, these proteins form the SARS-CoV-2 proteome [39].

The entry of the virus to the host cell is facilitated by the interaction of spike protein (S) with the host angiotensin-converting enzyme 2 (ACE2) receptor [2]. ACE2 receptors are located on the surface of epithelial cells. The epithelial cells constitute the outer most layer of most organs including the blood vessels and the lungs [7]. The density and distribution of ACE2 provide indication of potential infection avenues for the virus [42]. Transmembrane protease/serine subfamily member 2 (TMPRSS2) is a human protease which enhances viral entry as well. This is achieved through priming of the (S) protein. The S protein consists of two sub units: S1 and S2. S1 has the receptor-binding domain (RBD). This RBD has amino acids that play an essential role in protein-receptor binding [37]. The entry of virus into the cells is facilitated by the conformational alterations of S2 sub-unit resulting from binding of RBD with the ACE2 receptor. TMPRSS2 inhibitors could thus be considered as possible therapeutic targets for drug development [27, 28].

Several mathematical models [3, 15, 21, 22, 26, 33] have been developed to provide insights in the transmission dynamics and control of COVID-19 infection. These models have been useful for decision making by health professionals, officers and governments on how to manage and control COVID-19. An in-depth understanding of the interactions of SARS COV-2 virus with the immune system has the potential to provide specific targets for drug development, inhibiting the propagation of the virus. We therefore propose a mathematical model to describe the course of SARS COV-2 virus. We analyze the importance of the immune cells on the virus and the virion producing cells and identify potential targets for drug development against the virus.

The rest of the paper is organised as follows. In Section 2, the life cycle and viral infection of SARS COV-2 virus is reviewed. In Section 3, a deterministic model is presented with detailed mathematical analysis. Numerical simulations are carried out in Section 4. Finally, Section 5 gives the conclusion and recommendations for future work.

## 2 SARS CoV-2 infection and immune response

### 2.1 Viral entry and release

SARS CoV-2 entry is facilitated by the binding of spike proteins to ACE2, located on the surface of epithelial cells [20]. These viruses invade the host cells (1) via the cells surface following activation by serine proteases such as TMPRSS2 or (2) via endocytosis within endosomal–lysosomal compartments including processing by lysosomal cathepsins. When TMPRSS2 is expressed the previous pathway is favourably chosen whereas in the absence of the protease, the latter pathway is preferred [24]. Studies in [30] have shown that the spike proteins in SARS CoV-2 undergoes cleavage through TMPRSS2 to allow activation [30]. The spike protein contain two subunits: *S1* subunit (globular domain) that codes for a receptor binding subunit that facilitates viral binding to ACE2 receptor and *S2* subunit (transmembrane domain) that ensures fusion with the host membrane [14].

Upon binding to the ACE2, the S protein is bisected by the TMPRSS2 that acts as the cleaving enzyme [20] [30]. Furin activates *S2* subunit to ensure fusion or viral protein in addition to enabling cleavage of spike protein. Following fusion, the viral mRNA enters the host cells to initiate infection. Once inside the host cell, the single-stranded positive sense RNA acts as messenger RNA (mRNA) [30]. It is further translated by the host ribosomes, leading to the production of new virons [31]. Viral RNA synthesis takes place in two stages: genome replication and subgenomic mRNAs transcription followed by viral translation and assembly of viral particles. The initial stages are mediated by the replication/transcription complex (RTC) [14].

Functional proteins from the N- to C-termini of polyprotein involved in the replication process include: RNA-dependent RNA polymerase (RdRp, Nsp12), the zinc-binding helicase (HEL, Nsp13) and those with enzymatic functions related to RNA modifications such as mRNA capping (Nsp14, Nsp16), RNA proofreading (Nsp14), and uridylate-specific endoribonuclease activity (NendoU, Nsp15) [19]. The activity of these enzymes is further regulated by the association with other non-structural proteins (Nsp7–Nsp10) which are probably necessary to achieve all of the replication and transcription processes. [38]. Following post mRNA modifications, virons are released from the infected cell via budding, exocytosis or cell death. For immature virion particles, budding occurs. During this event, the N protein, whose primary role is to allow proper alignment for virion budding, interacts with host membrane and enable the release. Fully developed viruses that have matured at the golgi apparatus or endoplasmic reticulum are released via exocytosis [30].

### 2.2 Immune System and Viral Infections

During respiratory viral infections due to SARS COV-2, a cascade of events take place via mediation of innate responses and the epithelial cells with *ACE*2 receptors. The detection and recognition of viral RNA by the endosomal PRRs results in activation of various transcription factors which in turn triggers the expression of type 1 and type 3 interferons. These molecules also alert the neighbouring epithelial cells of an invasion. The normal cells stimulate anti-viral immunity to prevent attack from the virus. [25].

The adaptive immune, in contrast, takes longer to be established [35]. In addition to its specificity it produces memory cells which provides immunity in case of re-infection. The adaptive system is mobilized when the pathogen overwhelms the innate system. The dendritic cells migrate to the lymph and stimulates the activation of the adaptive system. The adaptive system is divided into cell-mediated immune response, which is carried out by T cells, and the humoral immune response, which is controlled by activated B cells and antibodies. Similarly, both T and B cells recognize the antigen via a complimentary receptor followed by self-maturation to specifically bind to the particular antigen of the infecting pathogen [35].

Naive T cells express either *CD*4^+^ or *CD*8^+^ cells [9]. Naive *CD*4^+^ cells binds to the antigen presenting cells (APC) to stimulate the maturation to T helper cells which stimulate the differentiation and maturation of B cells which secretes cytokines. Naive *CD*8^+^ cells differentiate to cytotoxic T cells which are directly involved in the eradication of the pathogens [9]. Recent investigation on middle aged COVID-19 patients [1] revealed the reduction in type 3 interferon(*IFNγ*) which is stimulated by the *CD*4^+^ cells. Striking difference appears where *CD*8^+^ cells have been shown to be hyper-activated and exhausted concurrently.

There are two categories of immune responses generated by immune system during viral infections: lytic and non-lytic mechanisms. The former involves killing of infected cells while the latter applies mechanisms to hinder viral replication. Cytotoxic T lymphocytes are highly involved in killing infected cells. Antibodies play a fundamental role in deactivating free virus resulting in inhibition of infected cells [5]. Over the years, coronaviruses have evolved and developed mechanisms to evade immune system. These mechanisms have been observed in the previous betacoronaviruses outbreaks and recent findings suggest SARS CoV-2 is no exception [11].

## 3 Mathematical Model

### 3.1 Model Formulation

To understand the within-host dynamics of SARS-CoV-2, we develop a deterministic model comprising the following compartments: uninfected epithelial cells (*E*_*S*_), latently infected epithelial cells (*E*_*IL*_), productively infected epithelial cells (*E*_*IP*_), free virus particles (*V*), and immune cells (*N*).

The model assumes that the uninfected epithelial cells, (*E*_*S*_), are generated at a constant rate Λ in the body with a carrying capacity of *K*_1_ and a natural death rate *µ*_1_. These cells become infected by the virus (*V*) at a rate modulated by immune inhibition, represented by a saturating function 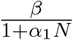 The infection results in the transition of the uninfected cells to latently infected cells, (*E*_*IL*_). At this stage, the virus attains a state of dormancy as it undergoes the various stages of viral infection including translation of the viral RNA. This leads to formation of enzymes that are involved in viral replication, and consequent generation and assembly of new viral particles. These dormant cells remain undetected by the hosts immune system through processes like down-regulation of the Major Histocompatibility Complex (MHC) and inhibiting the apoptotic pathway but can die naturally at a rate of *µ*_2_.

Subsequent to the dormancy state, the infected cell progresses to a productive state (*E*_*IP*_) which involves viral shedding. Similarly, this progression is limited by the interaction with the host immune response hence represented as 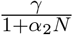 where *γ* is the progression rate and *α*_2_ represents immune inhibition. Virion release from the infected cells occur primarily through budding, exocytosis or cell death. In contrast to dormant infected cells, these productive cells are recognized by the immune system and mechanisms are initiated to destroy them. The number of virions released by the death of productive infected cells is represented by *κ*. The virions die naturally or due to destruction by the immune cells at a rates *δ* and *v*, respectively. The immune cells are produced at a rate Π with a carrying capacity *K*_2_. Following a viral infection, Immune cells are activated proportionally to antigen presence with saturation, modeled as (*ϵNV*)*/*(*ζ* + *V*), where *ϵN* is the maximum rate of immune cells production and *ζ* depicts when the rate of interaction between the immune cells and virus is half the maximum rate. The immune cells die naturally at a rate *σ*.

Additional model assumptions include:

- The SARS COV-2 selectively attacks the epithelial cells with *ACE*2 receptors.
- The uninfected epithelial cells, *E*_*S*_ are constantly being produced in the body until they reach the carrying capacity.
- There exists natural death rates for cells and virions.
- The latent cells are undetected by the immune system and any interventions directed to these cells are non-lytic, limiting viral replication.
- Production of the virons occur solely due to lysis of the infected cells.
- The virus levels induce the production of the immune response cells. The increase in response is assumed to be proportional to the level of virions present.
- The model does not differentiate between the various components of the immune system such as cytotxic T-cells and antibodies.

A summary of the parameters and variables are provided in Table 1 and Table 2, respectively.

**Table 1.**
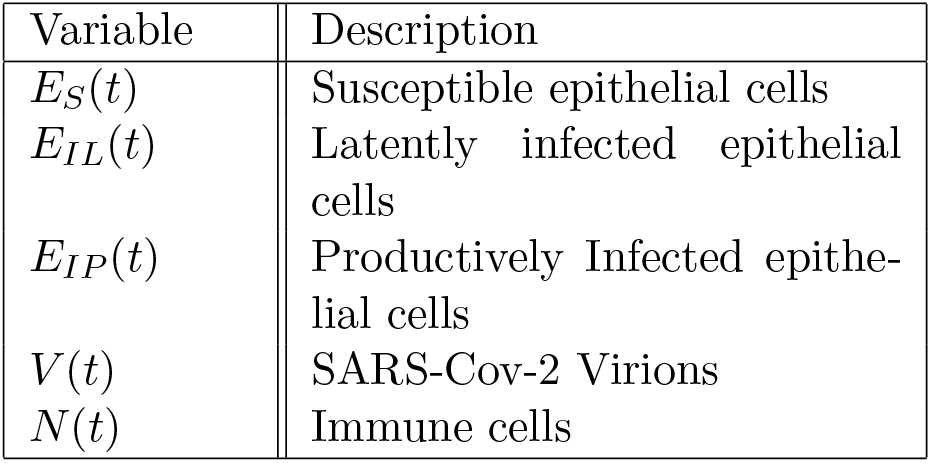
Description of model variables.

**Table 2.** Description of model parameters.

| Parameter | Description |
| --- | --- |
| $\Lambda$ | Recruitment rate of susceptible epithelial cells |
| $K_1$ | Carrying capacity of the epithelial cells |
| $K_2$ | Carrying capacity of the immune cells |
| $\mu$ | Natural death rate of epithelial cells |
| $\delta$ | Natural death rate of virus |
| $\sigma$ | Natural death rate of immune cells |
| $\beta$ | Infection transmission rate |
| $\rho$ | Removal rate of the infected epithelial cells due to interaction with immune cells |
| $\Pi$ | Number of immune cells produced |
| $\nu$ | Removal rate of the virus due to interaction with immune cells |
| $\kappa$ | Number of virions produced from infected cells |
| $\gamma$ | Progression rate of latently infected epithelial cells |
| $\alpha_1$ | Intervention rate by the immune cells to virus infection |
| $\alpha_2$ | Intervention rate by the immune cells to transition of latent epithelial cells |
| $\zeta$ | Michaelis-Menten constant-measure of cytokines production |
| $\epsilon$ | Recruitment rate of immune cells due to viral invasion |

A compartmental representation of the SARS CoV-2 *in-vivo* dynamics is shown in Figure 1.

**Figure 1.**
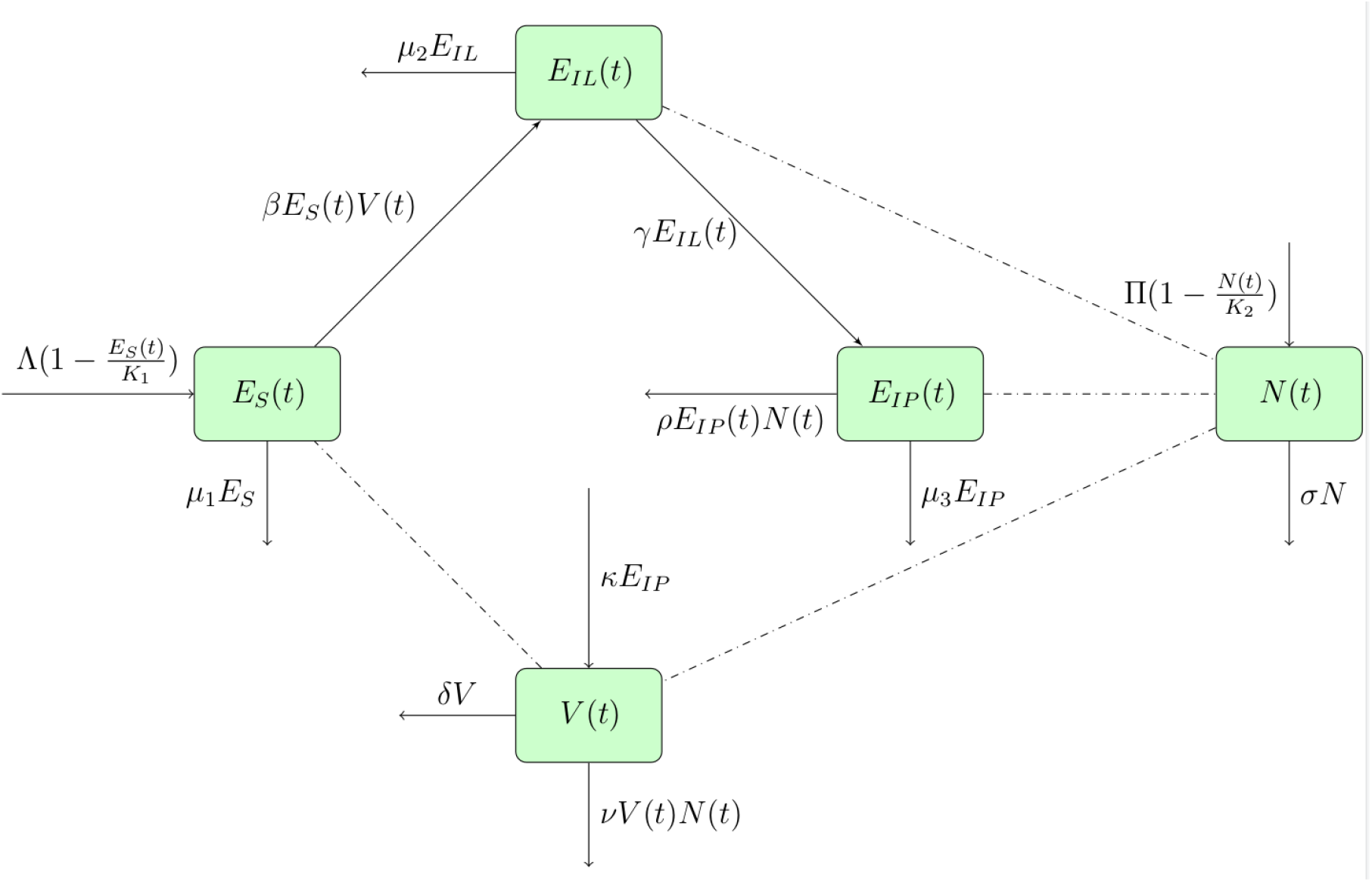
A compartmental representation of the SARS CoV-2 *in-vivo* dynamics.

Based on the above model description and assumptions, a deterministic model for SARS CoV-2 in-vivo infection dynamics is presented as follows:

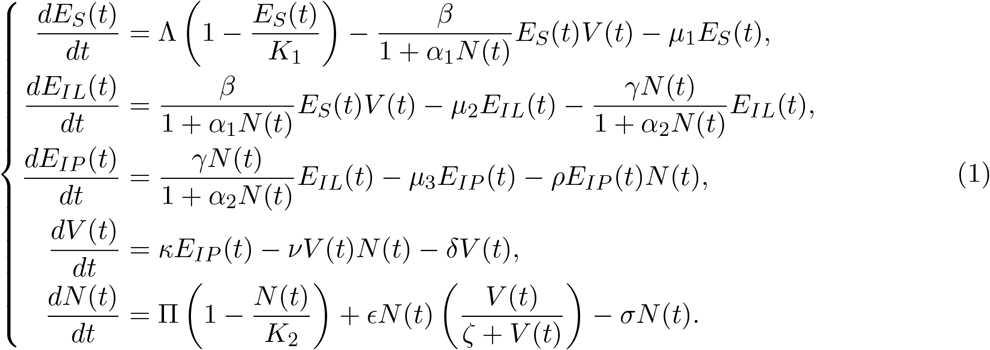

where *E*_*S*_(0) *>* 0, *E*_*IL*_(0) *≥* 0, *E*_*IP*_ (0) *≥* 0, *V* (0) *≥* 0, *N* (0) *>* 0.

### 3.2 Model Analysis

#### 3.2.1 Positivity of Solutions

All state variables (*E*_*s*_, *E*_*IL*_, *E*_*IP*_ , *V, N*) are non-negative and that the solution of the system (1) with positive initial values remain positive for all *t ≥* 0.

From the first equation in system (1):

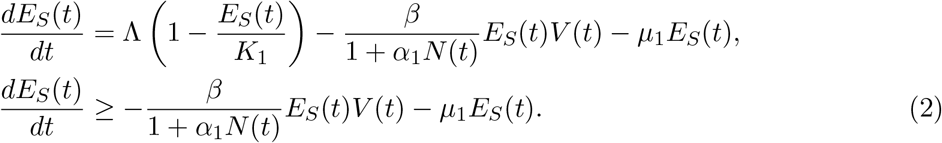

Assuming no contribution from the immune system, equation (2) becomes:

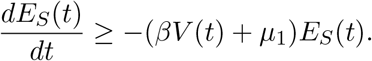

Using separation of variables and applying initial conditions in system (1),

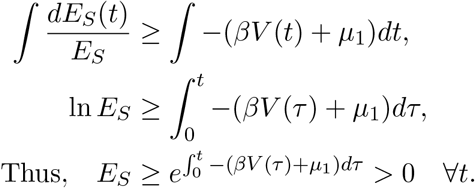

Using a similar approach, the rest of the equations in system (1), can be presented as follows:

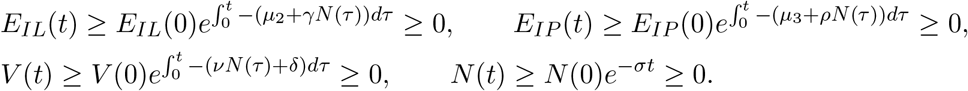

Hence, the state variables (*E*_*s*_, *E*_*IL*_, *E*_*IP*_ , *V, N*) are non-negative ∀t > 0.

#### 3.2.2 Boundedness of the Solutions

Let the total population of epithelial cells be *E*_*T*_ , where *E*_*T*_ = *E*_*S*_ + *E*_*IL*_ + *E*_*IP*_ . Substituting the derivatives in system (1), we have

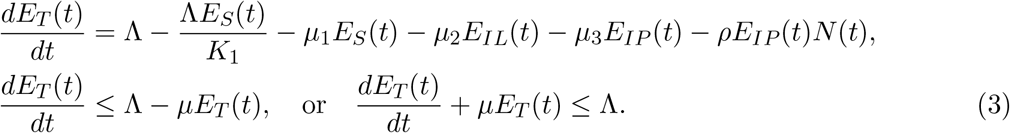

Upon integrating equation (3) and applying the initial condition in system (1), we obtain

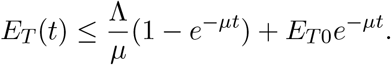

Clearly, as *t → ∞*, 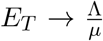 That is, there exists a bounded positive invariant region for the epithelial cells. Thus,

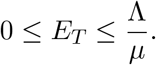

Similarly, for the immune cells *N* (*t*), we have:

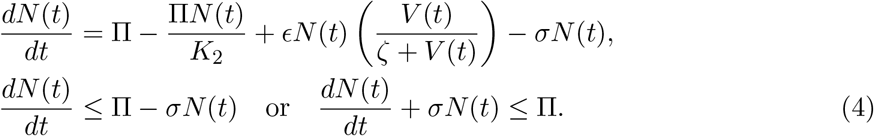

Solving equation (4) yields:

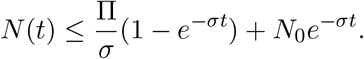

Clearly, there exists a bounded positive invariant region for the immune cells. That is,

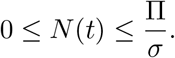

This yields the following Lemma.

**Lemma 1**. *The feasible region* Ψ *defined by*

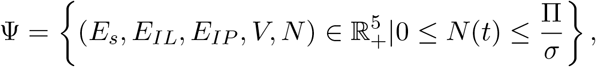

*with the non-negative initial conditions E*_*s*_(0) *>* 0, *E*_*IL*_ *≥* 0,*E*_*IP*_ *≥* 0, *V ≥* 0, *N >* 0, *is positively invariant*.

### 3.3 Equilibrium Analysis of the Model

The model system (1) has a disease-free equilibrium point, denoted as *E*_0_ where

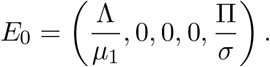

Using the next generation matrix approach, the basic reproduction number, *R*_0_ for the disease was computed. *R*_0_ = *ρ*(*FV* ^*−*1^), where *ρ* is the spectral radius of the next generation matrix *FV* ^*−*1^. *F* is the matrix of new infections while *V* is the matrix of transfer of infections from one compartment to the other.

From the Jacobian matrix of system (1), we obtain

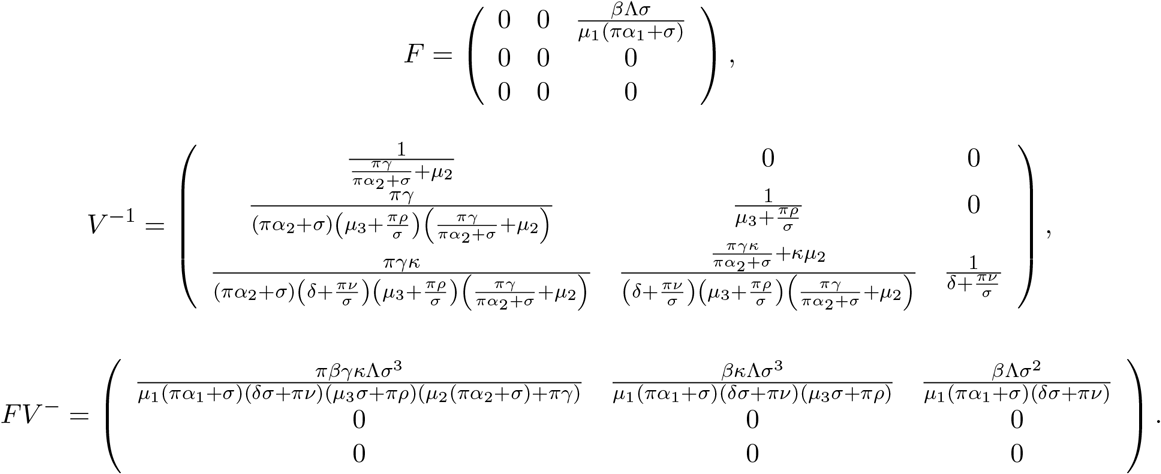

Therefore,

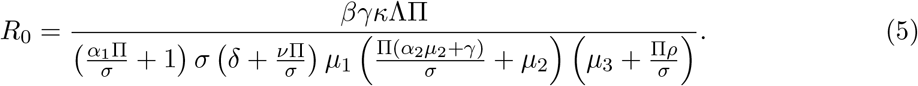

*R*_0_, given by (5), describes the average number of newly infected cells produced by an infected cell during its infectious period. Hence, this ratio determines whether the virus persists within the host or becomes extinct. If *R*_0_ is above unity, an infection is initiated.

#### 3.3.1 Local stability of the disease-free equilibrium point, *E*_0_

The disease-free equilibrium *E*_0_ is locally asymptotically stable if the eigenvalues of the Jacobian matrix of model system (1) are negative or have negative real parts (if complex). The Jacobian matrix of system (1) is given as follows:

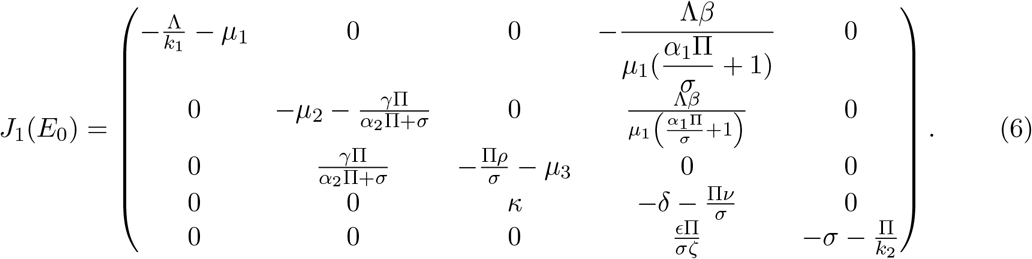

From the first column of matrix (6), we have 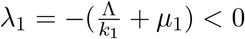 Upon deleting the first row and first column, the matrix (6) is reduced to:

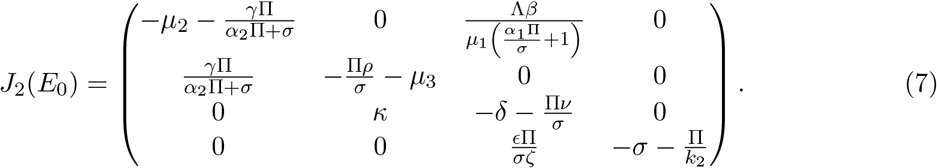

Again, from column four in (7), we get 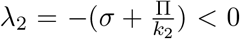 We further reduce matrix (7) by deleting the fourth row and fourth column to obtain:

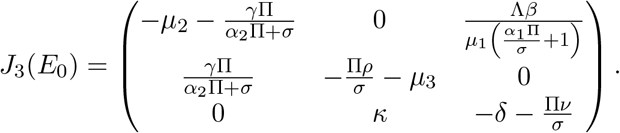

Matrix *J*_3_ has one negative real eigenvalue *λ*_3_ and a pair of complex eigenvalues with negative real parts, *λ*_4,5_ given as follows.

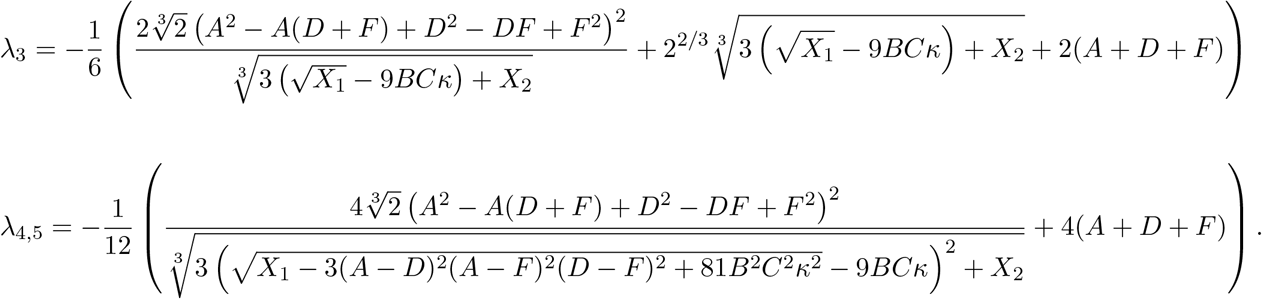

Where

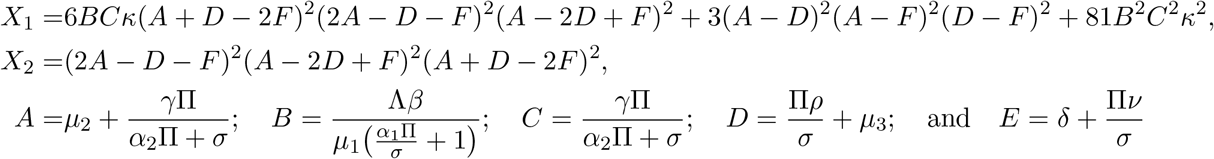

Clearly, all the eigenvalues of the Jacobian matrix in equation (6) are real and negative or complex with negative real parts. Hence, the model disease free equilibrium *E*_0_ is locally asymptotically stable.

#### 3.3.2 Global asymptotic stability of the disease-free equilibrium point

Following presentations in [16], we show that the disease-free state *E*_0_ is globally asymptotically stable when the basic reproduction number, *R*_0_ *<* 1. Rewriting the model system (1) in pseudotriangular form yields:

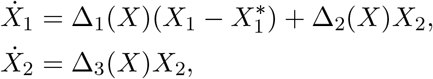

where *X*_1_ is the vector representing the non-transmitting states (susceptible epithelial cells *E*_*s*_ and immune cells *N*). The populations in these compartments are not infected with SARS-Cov-2 virus and not therefore transmit the viral infection. Thus, *X*_1_ = (*E*_*s*_, *N*). The vector *X*_2_ represents the compartments that are responsible for virus transmission within the human host. These states are infected or infective. Thus, *X*_2_ = (*E*_*IL*_, *E*_*IP*_ , *V*). We now have:

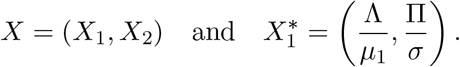

Linearization of model system (1) and subsequent computation of subsystem *X*_1_ yields

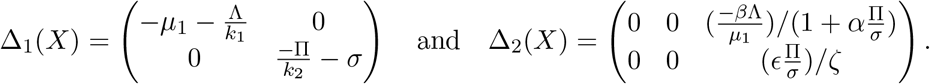

Clearly, the eigenvalue of matrix *Δ*_1_(*X*) are real and negative, showing that the system 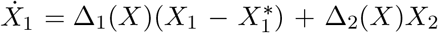 is globally asymptotically stable at *E*_0_. On the other hand, subsystem *X*_2_ yields matrix *Δ*_3_(*X*) as shown below.

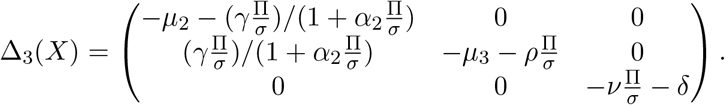

It can clearly be seen that *Δ*_3_(*X*) is a Metzler matrix i.e., all its off-diagonal elements are non-negative.

**Lemma 2**. *The Metzler matrix Δ*_3_(*X*) *is Hurwitz stable if and only if it is invertible and the matrix* 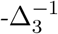 *is non-negative*.

In this case, *Δ*_3_(*X*) is a square matrix with a non-zero determinant given as

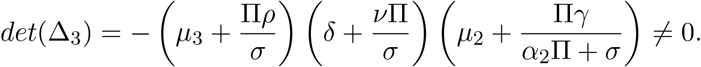

Hence, matrix *Δ*_3_(*X*) is invertible. Similarly, 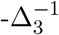 is given by

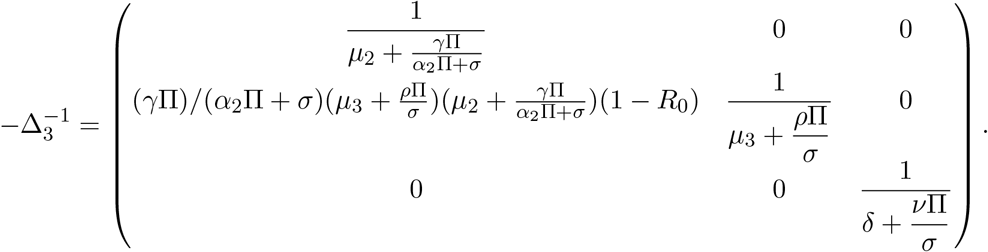

Observe that the 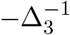 is non-negative when *R*_0_ *<* 1. Therefore, the disease-free equilibrium *E*_0_ is globally asymptotically stable when *R*_0_ *<* 1.

### 3.4 Existence of endemic equilibrium

The endemic equilibrium point, *E*_1_, of model (1) is calculated and given as

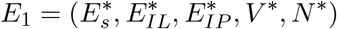

Where

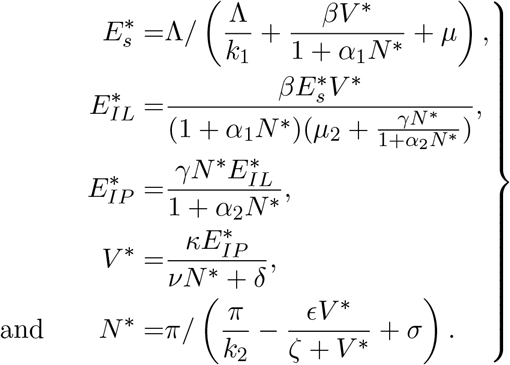

## 4 Numerical Simulation

### 4.1 Parameter Estimation

Limitations arising from experimental data on *in-vivo* dynamics of SARS CoV-2 resulted in use of parameter values from literature. Other values were assumed based on their biological feasibility. Table 3 summarizes all the parameter values.

**Table 3.** Parameter values for invivo COVID-19.

| Parameter | Value | Range | Source |
| --- | --- | --- | --- |
| $\Pi$ | $1 \times 10^4$ | $1 \times 10^4 - 1 \times 10^5$ | Assumed |
| $\beta$ | 0.0072 | 0-1 | [12] |
| $\gamma$ | 0.2 | 0-1 | Assumed |
| $\kappa$ | 500 | 88-580 | [12] |
| $\Lambda$ | 5000 | 57.757 - 12000 | [12] |
| $\mu_3$ | 0.4 | 0.088-0.58 | [12] |
| $\sigma$ | 0.67 | 0.05-1 | [12] |
| $\mu_1$ | 0.01 | 0-1 | Assumed |
| $\alpha_1$ | 0.000574 | 0-1 | [12] |
| $\delta$ | 0.8 | 0-1 | Assumed |
| $\nu$ | 0.6 | 0-1 | Assumed |
| $\rho$ | 0.00119 | 0-1 | [17] |
| $\alpha_2$ | 0.03 | 0-1 | Assumed |
| $\mu_2$ | 0.33 | 0.088-0.58 | [12] |
| $\epsilon$ | 0.1 | 0-1 | [12] |
| $\zeta$ | $1.26 \times 10^5$ | $1 \times 10^3 - 7.94 \times 10^7$ | [8] |
| $K_1$ | $4 \times 10^7$ | $1.27 \times 10^7 - 4 \times 10^8$ | [8, 12] |
| $K_2$ | $1 \times 10^7$ | $1 \times 10^7 - 1 \times 10^8$ | Assumed |

### 4.2 Sensitivity analysis of model parameters

Using the normalised forward sensitivity approach and the parameter values in Table 3, we computed the sensitivity indices of the different model parameters against the disease *R*_0_. Parameters with positive indices increases model *R*_0_ when they are increased. On the other hand, parameters with negative indices decreases the model *R*_0_ when they are increased. Results of sensitivity analysis are displayed in Figure 2 and Table 4.

**Table 4.**
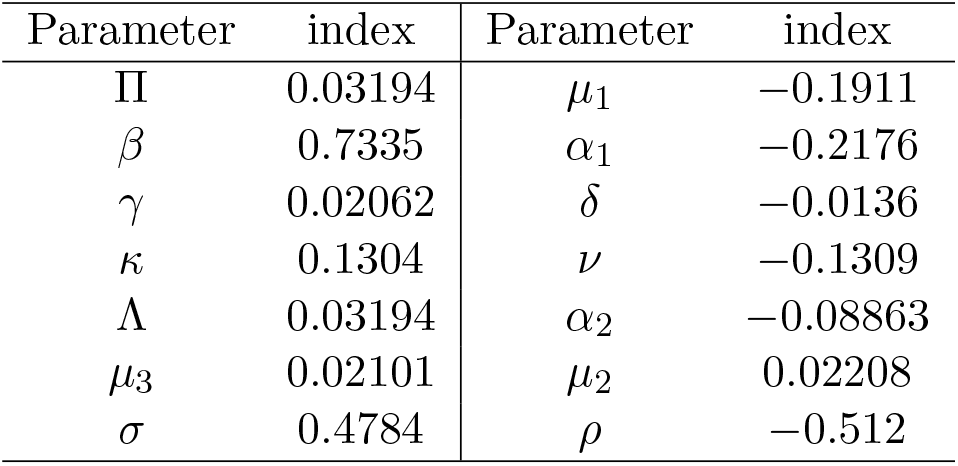
Sensitivity indices of Parameters.

**Figure 2.**
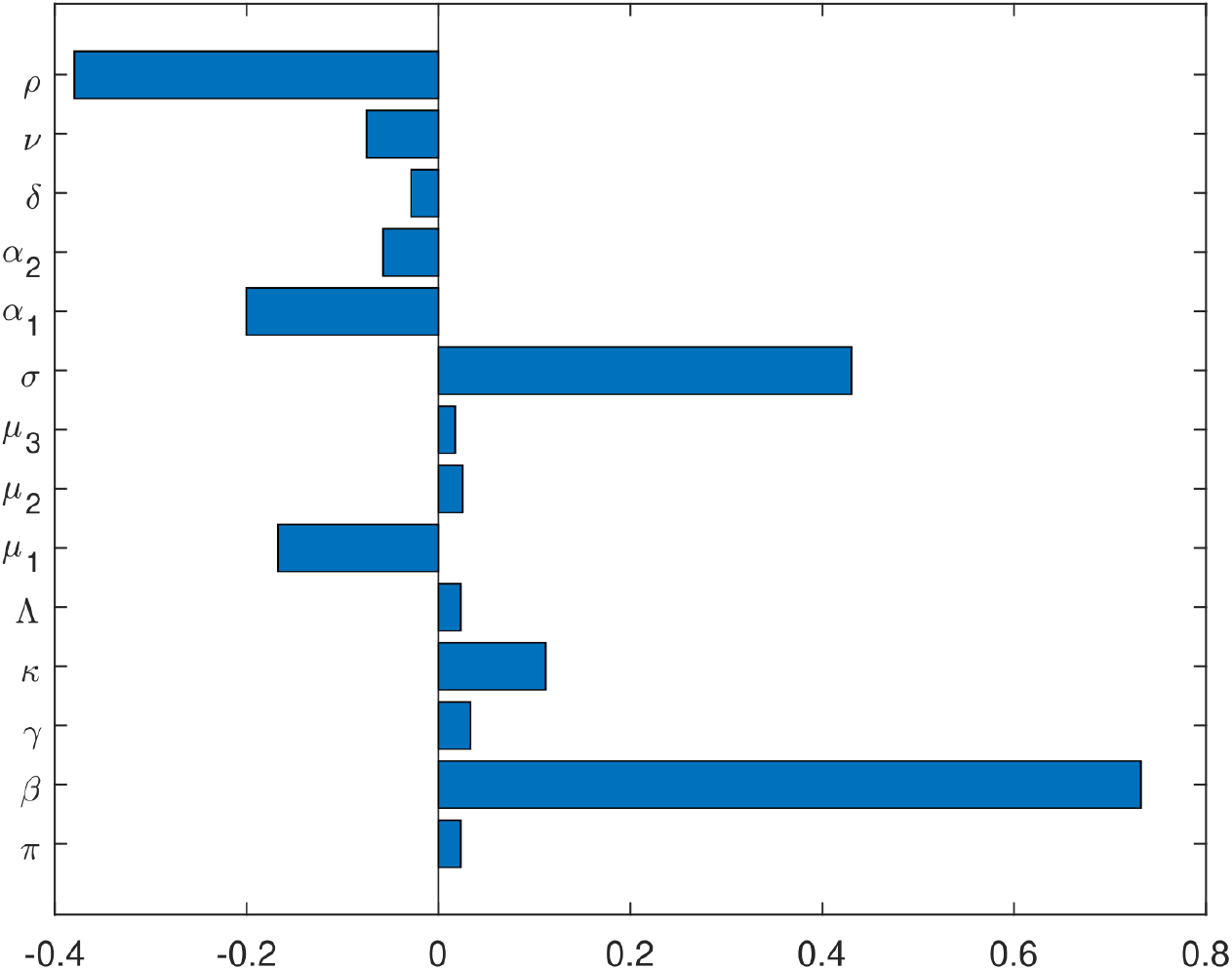
Tornado plots of in-vivo COVID-19

#### 4.2.1 Implications of the *R*_0_ Sensitivity Analysis

In the within-host context, the basic reproduction number *R*_0_ represents the average number of newly infected cells generated from one infected cell at the start of infection, when target cells and immune resources are abundant. Sensitivity analysis of *R*_0_ highlights the parameters with the greatest influence on viral persistence or clearance. The analysis indicates that increases in the infection rate (*β*) or the virus production rate (*κ*) drive *R*_0_ above the threshold value of 1, leading to rapid viral expansion. In contrast, increasing immune-mediated clearance rates (*α*_1_, *ρ, v*) or the natural decay rate of the virus (*δ*) can reduce *R*_0_, often pushing it below 1, resulting in viral decline [34].

From a therapeutic perspective, parameters with the highest sensitivity indices to *R*_0_ are prime drug or vaccine targets. Reducing *β* corresponds to blocking viral entry, as achieved by neutralizing antibodies induced by vaccines such as Pfizer-BioNTech, Moderna, and Oxford-AstraZeneca, or by fusion inhibitors. Lowering *κ* aligns with replication-blocking antivirals such as Remdesivir and Molnupiravir. Increasing *v* mimics enhancing viral clearance through immune-boosting vaccination or monoclonal antibody therapy.

### 4.3 Simulation Results

#### 4.3.1 Effect of Basic Reproduction Number on SARS-Cov-2 Viral load

When *R*_0_ *>* 1, the virus rapidly spreads within the host see red graph in Figure 3, depleting epithelial cells and sharply increasing virion count—this mimics the acute phase of infection. This could be as a result of lack of vaccination and/or comorbities among the hosts. At these levels, we would expect the infection to be severe often associated with abnormal increase in creatinine and lactose dehydrogenase levels, signalling kidney and livers dysfunctions respectively. Further, a low lymphocyte count is the most prevalent marker for SARS CoV-2 severity among unusually low haemoglobin levels and increased ferritin levels [6, 29]. Conversely, when *R*_0_ *<* 1, the immune system or intervention prevents persistent infection, and tissue health stabilizes as shown by the green line in Figure 3. This suggests that the infected individual has built enough immunity to recover from the virus without interventions such as vaccines.

**Figure 3.**
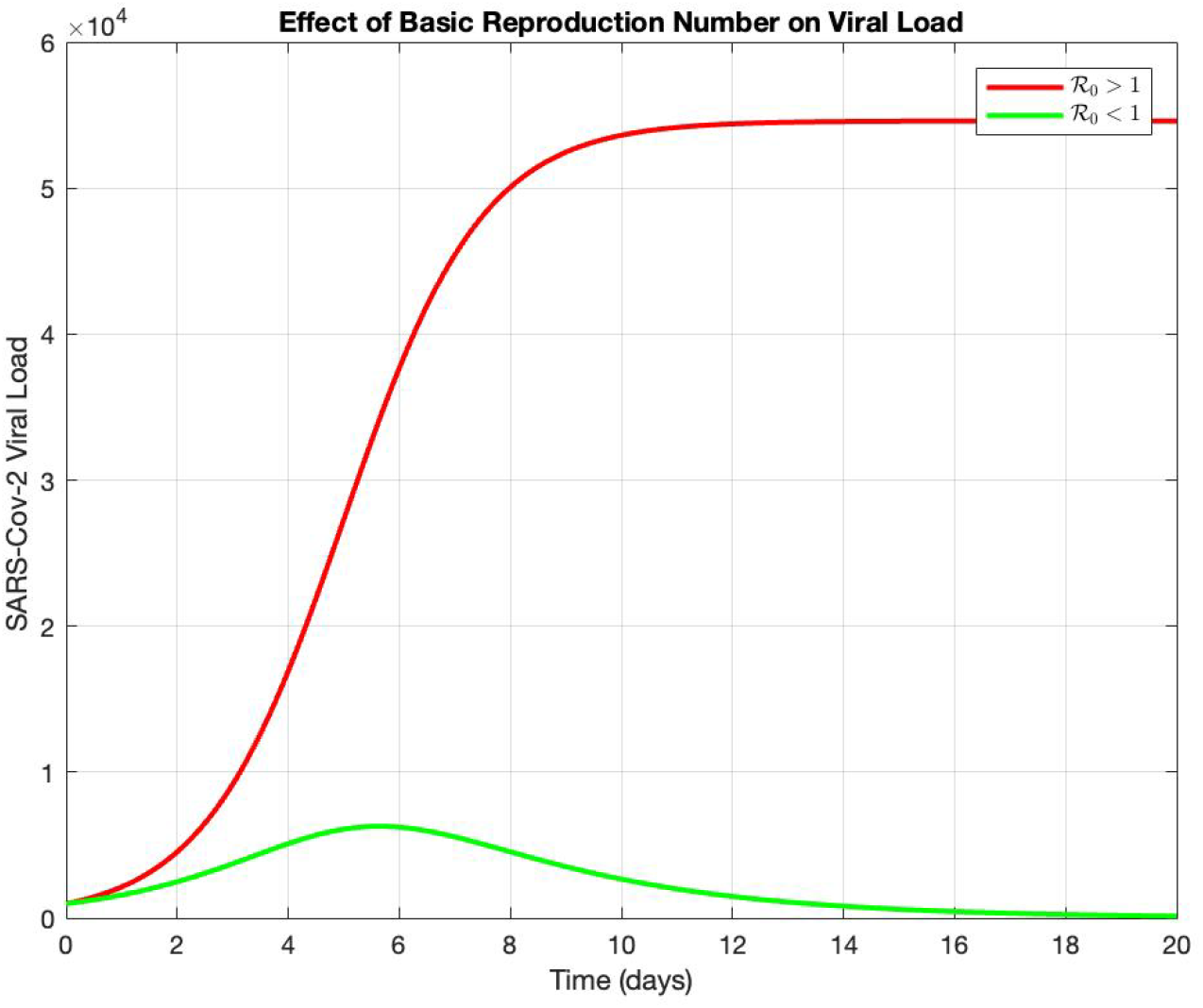
Effect of Basic Reproduction Number on Viral Load

The results indicate that combination strategies targeting multiple high-impact parameters are more likely to maintain *R*_0_ *<* 1 and achieve viral clearance. Furthermore, vaccines have the dual advantage of lowering both *β* and *κ* by preventing viral entry and promoting cytotoxic T-cell clearance of infected cells. This dual mechanism supports the preemptive administration of vaccines, which can reduce *R*_0_ before infection becomes established, ultimately limiting disease severity and transmission potential.

#### 4.3.2 Effect of Immune Efficacy on Viral load

Figure 4 compares three amplifications of immune function—entry blocking (*α*_1_), cytolytic activity against infected cells (*ρ*), and immune-mediated clearance of free virions (*v*) and their consequences for within-host viral kinetics. Although the plotted curves are schematic exponentials chosen to illustrate qualitative trends rather than direct ODE outputs, they align with the model mechanisms. The simulation comparing increases in immune-efficacy parameters (*α*_1_, *ρ*, and *v*) demonstrates how different immune mechanisms shape within-host viral dynamics.

**Figure 4.**
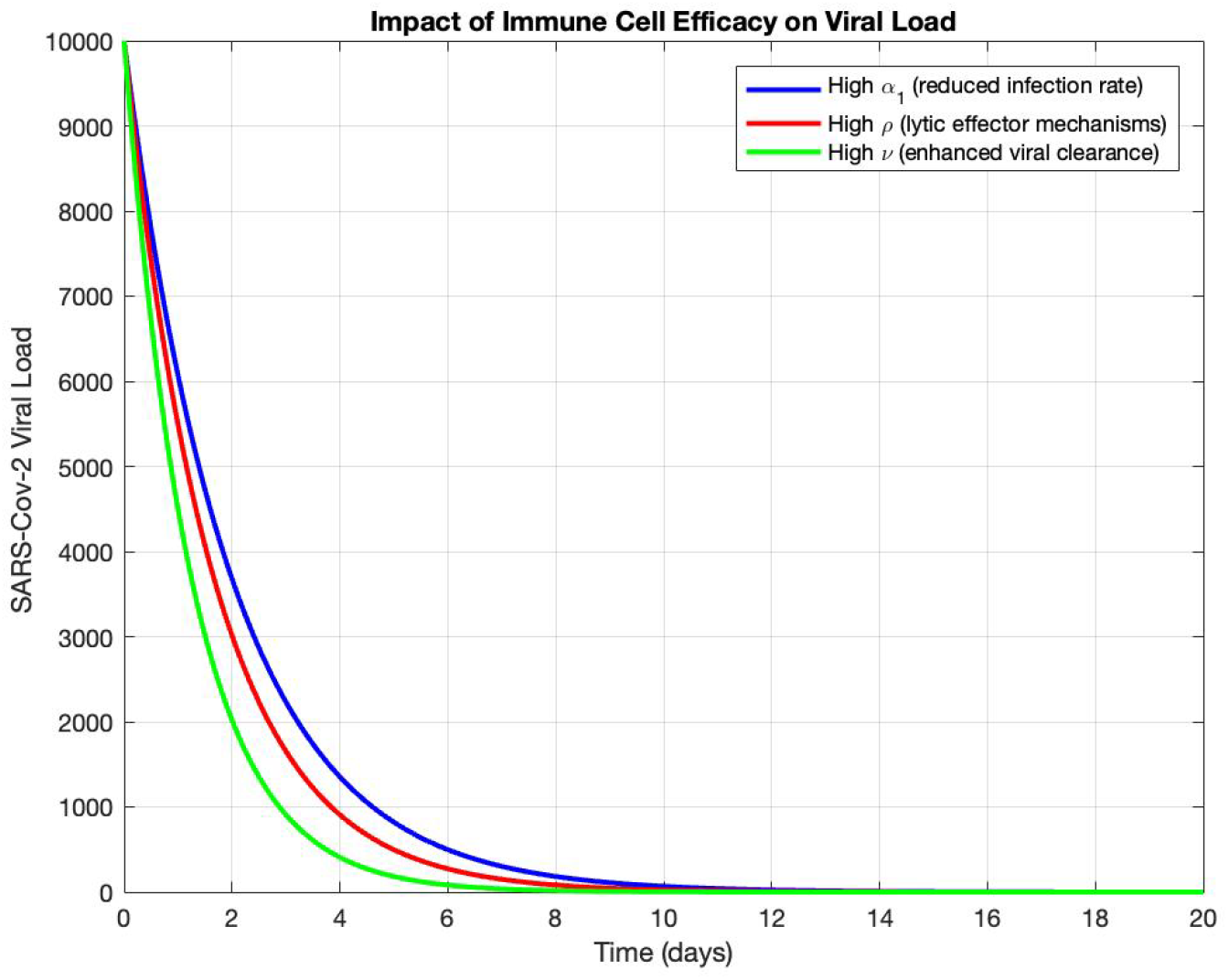
Effect of immune efficacy on viral dynamics

**Entry blocking (***α*_1_ *↑*). Increasing *α*_1_ reduces the effective infection rate (*β/*(1 + *α*_1_*N*)), slowing the early expansion of infection. The result is a lower and later viral peak because fewer susceptible epithelial cells become infected per unit time. This mechanism chiefly shapes the *establishment* phase of infection and is analogous to neutralizing antibodies or vaccines that impede cell entry.

**Cytolytic control (***ρ ↑*). Enhancing *ρ* accelerates removal of productively infected cells (*ρNEI*), which shortens the duration of productive infection and curtails ongoing virion production. Compared with *α*_1_, raising *ρ* has stronger effects after infection is seeded, reducing the plateau/shoulder of the curve and bringing the peak forward in time.

**Viral clearance (***v ↑*). Boosting *v* increases immune-mediated clearance of extracellular virions (*vNV*). In the simulations, this produces the steepest post-peak decline as shown by green line in Figure 4, reflecting rapid *debulking* of circulating virus. Because *v* acts directly on *V* , it yields the most immediate reduction in measured viral load among the three levers.

#### Discussion of the Impact of Immune Efficacy

Quantitative within-host simulations show that the three immune mechanisms act in complementary ways: *α*_1_ limits initial infection (seeding), *ρ* eliminates infected cells (intracellular factories), and *v* clears free virions (extracellular pool). Lytic effector responses, primarily mediated by cytotoxic T cells, increase the death rate of infected cells, while non-lytic mechanisms, such as neutralizing antibodies, block viral entry and replication [5, 41].

Increasing any of *α*_1_, *ρ*, or *v* suppresses viral load and reduces infected cell counts [10, 32], but the greatest benefit comes from combining these mechanisms. Such synergy lowers the peak viral load, shortens the infection duration, and reduces the overall viral burden (AUC). Clinically, this corresponds to interventions combining entry-blocking antibodies, cytotoxic T cell–mediated killing, and Fc-effector or innate clearance functions.

From a policy and clinical perspective, individuals with impaired immune efficacy (e.g., older adults or immunocompromised patients) are likely to experience higher viral peaks and slower clearance, underscoring the value of early, multi-mechanism strategies for both disease control and transmission reduction.

#### 4.3.3 Different Vaccine efficacy Impact on Viral load

The simulation compares two conceptual SARS-CoV-2 vaccine strategies: those that block viral entry (fusion inhibitors, represented here by vaccines that neutralize spike-mediated cell entry) and those that boost immune-mediated viral clearance (analogous to transcriptase inhibitors or immune-enhancing vaccines). Reducing *β* corresponds to blocking viral entry, as achieved by neutralizing antibodies induced by vaccines such as Pfizer-BioNTech, Moderna, and Oxford-AstraZeneca, or by fusion inhibitors . Increasing *v* mimics enhancing viral clearance through immune-boosting vaccination or monoclonal antibody therapy: more effective in long-term control as shown in red line in Figure 5a. This supports a therapeutic focus on intracellular targets (e.g., Remdesivir) rather than only blocking entry.

**Figure 5.**
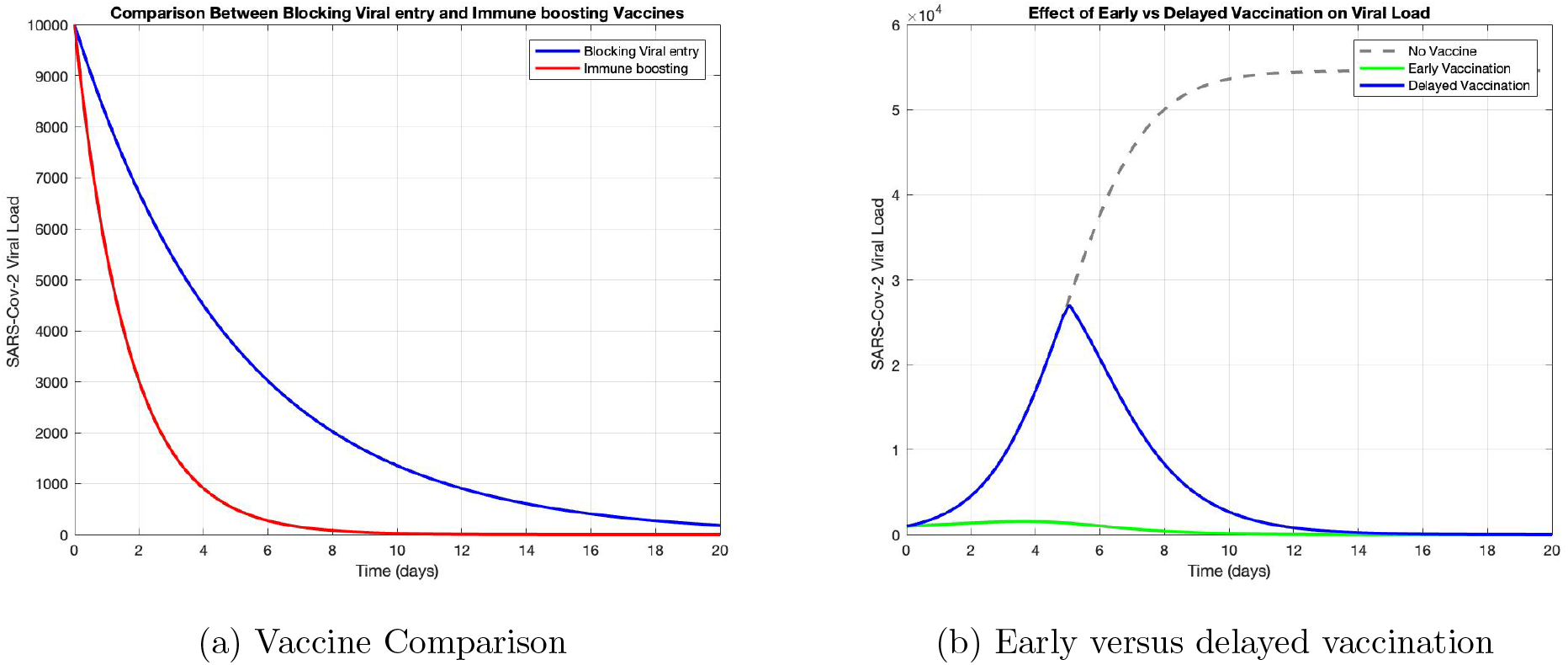
vaccination Impact

In Figure 5b, Gray dashed line (No vaccine) shows high, sustained viral load. Green (Early vaccination) shows viral load suppressed early. Blue (Delayed vaccination) shows partial control but delayed effect.

#### Discussion of the Impact Vaccine efficacy

Blocking viral entry produces a slower decline in viral load, reflecting the prevention of new infections but with limited immediate impact on virus already present in circulation. In contrast, immune-boosting approaches accelerate viral decay by enhancing immune clearance mechanisms, leading to a steeper decline and shorter infection duration. In comparing vaccine efficaciousness, immune-boosting vaccines outperform blocking entry therapies. The former block progression to virion-producing states, reducing viral burden even after initial infection. Fusion inhibitors prevent new infections but may not clear already infected cells. Also early vaccination controls the virus before it peaks. Delayed vaccination slows viral decline but can’t prevent peak damage. Overall, simulations emphasize that successful treatment must both prevent new infections and suppress intracellular replication.

#### 4.3.4 Vaccine and Immune response Impact on Viral load

SARS CoV-2 is a highly infectious disease caused by combined transmission from asymptomatic and symptomatic individuals [36]. SARS CoV-2 vaccines induce memory T and B-cells to combat future attacks and consequently boost immunity [4]. The simulation compares viral load trajectories under three scenarios: no vaccination, vaccination in a host with strong immunity, and vaccination in a host with weak immunity. In the absence of vaccination, viral load rises rapidly to a high peak before declining, as shown by Gray dashed line in Figure 6, reflecting uncontrolled replication until natural immune responses take effect.

**Figure 6.**
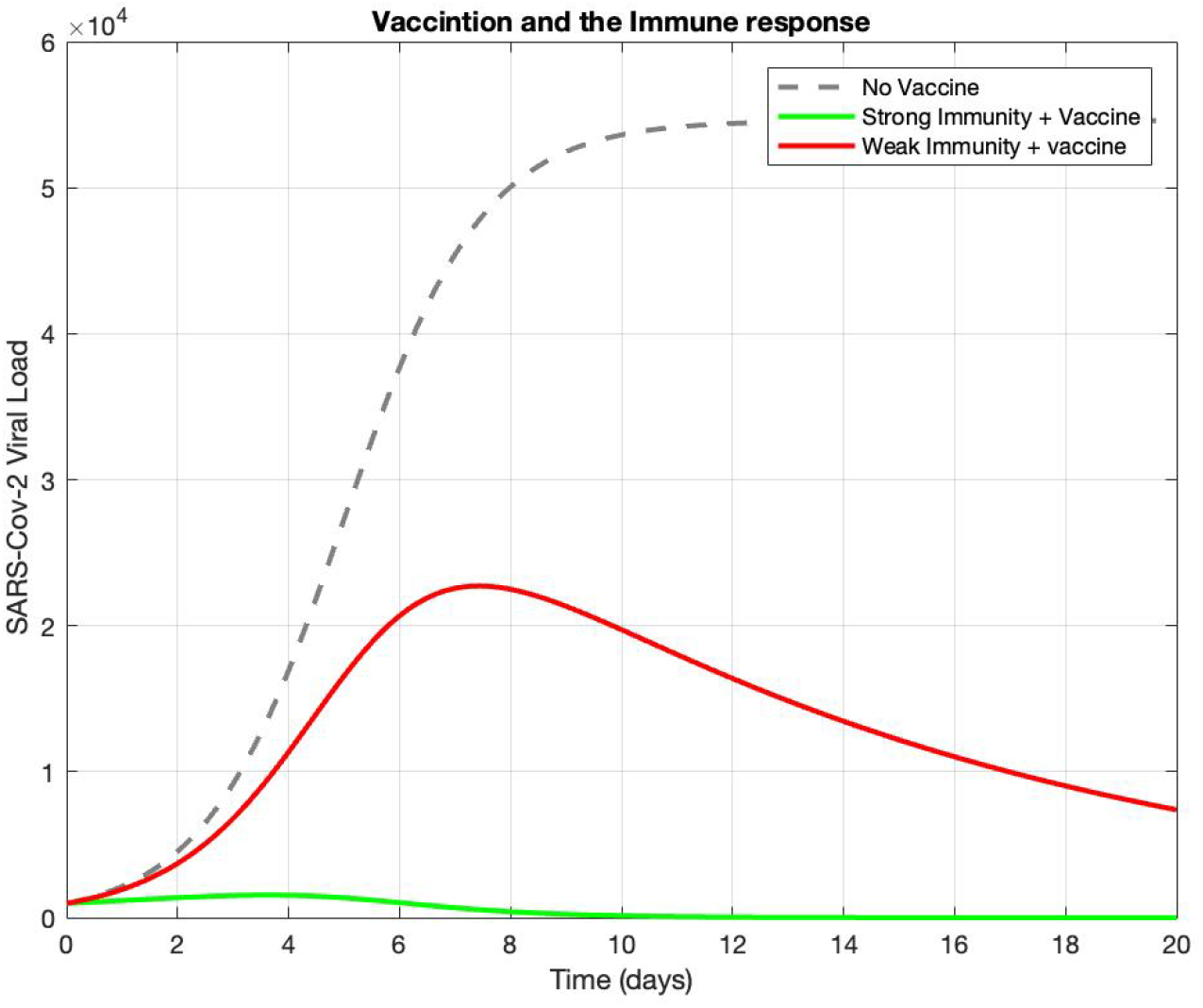
Vaccination and the Immune response

When vaccination is paired with strong baseline immunity, viral load is suppressed early, as indicated by the solid green line in Figure 6, with both the peak magnitude and the duration of infection substantially reduced. This reflects the combined effect of vaccine-induced protection and robust host immune clearance, preventing high viral titers from ever establishing.

In contrast, vaccination in a host with weak immunity shows a slower viral decline and a higher peak compared to the strong-immunity case as shown by red line in Figure 6, though still significantly better than no vaccination. This indicates that while vaccines provide protection in immunocompromised individuals, their effect is diminished without strong endogenous immune responses.

#### Discussion of Combination Therapy Under Varying Immune Strength

Enhancing immune efficacy (through higher *α*_1_, *ρ, v*) results in faster viral clearance and preservation of epithelial cells. These effects are amplified with vaccination, which promotes early and sustained immune activation.

From a clinical perspective, these results reinforce the importance of boosting immunity through prior vaccination, heterologous boosters, or immune-modulating interventions in vulnerable populations. Combination strategies that pair vaccination with therapies enhancing immune cell function may offer the greatest benefit for reducing viral burden and accelerating clearance.

## 5 Conclusion and Final Discussion

This study introduces and analyzes a mathematical model describing SARS-CoV-2 viral-host interactions within a human host. We rigorously explored the impact of immune responses and antiviral interventions on viral replication and tissue damage.

Our findings indicate that:

- The basic reproduction number *R*_0_ determines whether infection persists or clears.
- Immune responses (both lytic and non-lytic) are essential in suppressing infection.
- Vaccination enhances immune function, reducing viral load and preventing epithelial depletion.
- Immune-boosting interventions or monoclonal antibody therapy, which target intracellular replication processes, show superior long-term control over fusion inhibitors(Entry-blocking interventions).
- Timing matters – Early initiation of vaccination or therapy maximizes impact by preventing the virus from reaching high titers. Delayed interventions, although still beneficial, allow higher viral loads to persist for longer, potentially increasing disease severity and transmission risk.
- Policy and clinical implications – The findings support prioritizing early, multi-mechanism interventions, particularly for vulnerable groups such as the elderly and immunocompromised.

The model provides a foundation for studying future interventions, including combination therapy and immune modulation. Future work could extend the model to include pharmacokinetics/pharmacodynamics of antiviral agents, as well as fitting the model to patient-specific clinical data to improve its predictive power.

## Data Availability

All data produced in the present work are contained in the manuscript

## Acknowledgement

The authors are very grateful to Strathmore University and to the anonymous reviewers for their careful reading and constructive comments.

## Availability of data and materials

All data generated or analysed in this study are included in this manuscript.

## Authors’ contributions

All authors contributed to the writing and approval of the final manuscript.

## Ethical approval and consent to participate

Not applicable.

## Consent for publication

Not applicable.

